# Development and Evaluation of Two Rapid Indigenous IgG-ELISA immobilized with ACE-2 Binding Peptides for Detection Neutralizing Antibodies Against SARS-CoV-2

**DOI:** 10.1101/2020.12.19.20248535

**Authors:** Bijon Kumar Sil, Nihad Adnan, Mumtarin Jannat Oishee, Tamanna Ali, Nowshin Jahan, Shahad Saif Khandker, Eiry Kobatake, Masayasu Mie, Mohib Ullah Khondoker, Md. Ahsanul Haq, Mohd. Raeed Jamiruddin

## Abstract

COVID-19 pandemic situation demands effective serological tests with a view to adopting and developing policy for disease management, determining protective immunity as well as for sero-epidemiological study. Our study aims to develop and evaluate two rapid in-house ELISA assays targeting neutralizing antibodies (IgG) against S1 subunit of spike in SARS-CoV-2 and Receptor Binding Domain (RBD), as well as comparative analysis with nucleocapsid (NCP) ELISA. The assays were conducted with 184 samples in three panels collected from 134 patients. Panel 1 and 2 consist of RT-PCR positive samples collected within two weeks and after two weeks of symptom onset, respectively. Negative samples are included in panel 3 from healthy donors and pre-pandemic dengue patients. The total assay time has been set 30 minutes for both of the ELISA assays. Results show that S1 and RBD ELISA demonstrates 73.68% and 84.21% sensitivities, respectively for samples collected within two weeks, whereas 100% sensitivities were achieved by both for samples that were collected after two weeks of the onset of symptoms. S1-ELISA shows 0% positivity to panel 3 while for RBD-ELISA the figure is 1%. A strong correlation (r_s_=0.804, p<0.0001)) has been observed between these two assays. When compared with NCP-ELISA, S1 slightly better correlation (r_s_=0.800, p<0.0001) than RBD (r_s_=0.740, p<0.0001). Our study suggests S1-ELISA as more sensitive one than the RBD or nucleocapsid ELISA during the later phase of infection, while for overall sero-monitoring RBD specific IgG ELISA is recommended. Moreover, non-reactivity to dengue emphasize the use of these assays for serosurveillance of COVID-19 in the dengue endemic regions.

**Highlights:**

- The total assay time of these assays are 30 minutes.
- Sensitivity of S1 specific IgG ELISA for samples tested within 14 days of disease presentation is 73.68% while RBD specific ELISA demonstrates a sensitivity of 84.21%,
- Both of the assays under investigation can successfully detect all the cases (100% sensitivity) if the samples are tested after 14 days of onset of diseases.
- Specificity of S1-ELISA assay is 100%, whereas RBD specific IgG ELISA is 99% specific.
- The assays can be employed in dengue-endemic countries
- Among the three in-house IgG ELISA, assay system specific to S1 is found to be more sensitive and specific for retrospective serosurveillance.
- For acute to late phase, as well as retrospective serosurveillance of COVID-19, RBD-ELISA can be a method of choice for SARS-CoV-2 prevalent areas.

## Introduction

The novel Severe Acute Respiratory Syndrome Coronavirus 2 (SARS-CoV-2) emerged in Wuhan, China in the late 2019, has already afflicted countries across the world with COVID-19 disease [1]. The pandemic has already affected 218 countries with more than 1.5 million deaths [2]. The clinical spectrum of COVID-19 is reported to vary from asymptomatic to mild (80.9% of total cases) and severe (14% of cases) or life-threatening requiring medical interventions [3]. The multiple transmission routes such as droplets and direct contact, facilitated enhanced transmission of this disease, resulted in higher number of total deaths than by either SARS or MERS [4-6].

Early diagnosis of the disease is the most plausible approach to combat COVID-19. The World Health Organization has recommended molecular tests specifically qRT-PCR as the mainstay of diagnostic repertoires [7]. However, molecular detection of viral RNA is feasible only at the acute phase of infection, thus limiting its implementation when acute symptomatic phase is escaped [8]. Decisions exclusively based on molecular diagnostics, for patient management, restrictions on travelling as well as management of disease are now considered to be inadequate and worthy of a second-thought [9]. Countries considering gradual lifting of restrictions in order to draw the balance between economic and social damage posed by COVID-19, may experience a resurge of cases [10]. The status of virus circulation and prevalence in the population and whether there is protection against re-infection needs to be evaluated and these factors underpin the inclination towards development and worldwide approval of serological tests.

Neutralizing antibodies provides protection against viral infection and contribute in clearance of viruses and hence used as immune product for treatment against viral diseases. In order to determine efficacy of vaccines against viral diseases, determining the neutralizing antibody titre is practised throughout the ages [11, 12]. In this pandemic situation, plasma transfusion from recovered patient is considered in many countries as potential and available therapeutic intervention to treat critical patients [13], which also requires evaluation of plasma for neutralizing antibodies, thus accentuate the need of effective serological assays.

SARS-CoV-2 consists of structural and non-structural proteins with 12 putative functional ORFs (Open Reading Frames) with 82% similarities of nucleotides of SARS-CoV. The notable structural proteins of SARS-CoV-2 include Spike (S), Envelope (E), Membrane (M), Nucleocapsid (N) [14]. The trimeric S protein is composed of S1 and S2 subunits, while S1 comprising the receptor binding domain (RBD) and S2 forming fusion peptides, play key role in viral entry and can be exploited as a potential neutralizing target like in MERS and SARS as well as a diagnostic tool [15-18].

Recently, several diagnostic kits have been manufactured and commercialized based on several proteins of SARS-CoV-2 targeting all or segment of either of Nucleocapsid (NCP), Spike (S) or Receptor Binding Domain (RBD) proteins [19]. Currently, twelve ELISA kits have been approved by FDA (data on December 4, 2020) detecting pan-Ig, IgG, IgM or IgA [20]. ELISA kits targeting neutralizing antibodies are yet inadequate to satisfy the surge. Our study focuses on using RBD and S1 as antigen to develop two ELISA assays detecting neutralizing antibodies. We evaluated and compared the performance of two ELISA assays with each other and also with an in-house NCP-ELISA assay.

## Materials and methods

### Reagents and materials

ACE-2 receptor binding recombinant spike 1 and RBD (receptor binding domain) protein specific to SARS-CoV-2 from Sino Biologicals, China, was used as capturing agent. For detection of immunocomplex of human IgG-capturing antigen commercially bought Goat anti-human IgG conjugated with HRP (horseradish peroxidase) (Native Antigen, UK) was utilized as secondary antibody. 3,3′,5,5′-Tetramethylbenzidine (TMB) (Dojindo Molecular Technologies, USA) was used as HRP substrate (Wako, Japan) and the colour developed by TMB-peroxidase was stopped with 1.5 M H_2_SO_4_ (Sigma-Aldrich, Germany). ELISA reaction was read at 450 nm using ELISA plate reader (Thermo-Fisher Scientific, USA). 96-well flat-bottom polystyrene micro-titter plate (Extra Gene, USA) was used for assay development as test medium.

### Specimen Selection and Assortment

In order to develop and evaluate two assay systems, System 1: SARS-CoV-2 spike 1 specific ELISA assay and system 2: SARS-CoV-2 RBD specific ELISA assay, serum samples (n=184) were collected from 134 individual. Positive samples (n=79) were from confirmed RT-PCR positive SARS-COV-2 infected individuals categorised into two different panels: i) SARS-COV-2 infected patients with less than 14 days from symptom onset (n=19) ii) SARS-COV-2 infected patients with more than 14 days from symptom onset (n=60). There was a third panel, consisted of negative samples including i) Samples collected from dengue positive individuals prior to the SARS-CoV-2 outbreak (n= 24) and ii) Samples collected from healthy individuals during April to June (n=81). All the samples were stored at −80°C until further use.

### Categorization of Seropositive and Seronegative samples

Positive panel and negative panel were characterized by four factors: i) clinical symptoms development, ii) Outcomes of RT-PCR test, iii) Outcomes based on a developed in-house ELISA assay targeting SARS-CoV-2 nucleocapsid protein (NCP), and iv) Pre-pandemic samples. Based on RT-PCR, in-house NCP ELISA and commercial Elecsys Anti-SARS-CoV-2 immunoassay, two positive control and two negative control sera were selected for development and optimisation [21].

### Optimization and standardization of ELISA systems

Checkerboard test (CBD) was applied for the standardization and optimization of test components of both S1 and RBD ELISA assays. Test conditions were determined by analysing multiple combinations of conditions using an in-house developed IgG ELISA (SARS-CoV-2 nucleocapsid) assay as reference [21] and adopted the most optimal conditions. In order to determine the optimal duration, different time span for antigen coating and blocking, sample incubation, conjugation and substrate exposure were assessed. The positive and negative sera were diluted at a range of 1:10 to 1:200 and tested against dilution range 1:100 to 1:600 and 1:1000 to 1:6000 for optimum concentration of SARS-CoV-2 proteins (S1 and RBD) and anti-IgG conjugate, respectively. From the multiple combinations, the condition that showed the best signal to noise ratio (S/N) with acceptable background was decided to be optimum and has been selected. The formula used for calculating the S/N was,

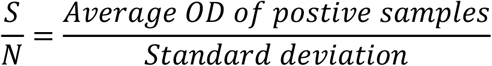

### Preparation of platforms for in-house ELISA

Briefly, two 96-well microtiter plates were coated with two separate commercial recombinant SARS-CoV-2 (S1 and RBD) antigens (100 μl/well), diluted in sodium-bicarbonate buffer (pH >10). The antigen-coated plates were either kept at 37 °C for an hour or overnight at 4 °C. Unbound antigens were washed off after immobilization prior to blocking (100 μl/ well), using phosphate-buffered solution (PBS) containing 0.01% tween and 2% BSA. ELISA plates were incubated at 37 °C for one hour. After incubation, ELISA plates were either used for subsequent steps or kept at 4 °C until use. Before use, the wells were cleared, followed by three washes with 300 μl/well in-house ELISA wash buffer (50 mm Tris, 0.05% Tween 20, 0.1% SDS, 0.8% NaCl, distilled water).

### Assay procedure optimisation

100 μl of human serum samples diluted in a dilution buffer (PBS, 0.1% Tween 20, 1% BSA) at 1:100, were added to the wells and incubated at 37°C for 15 minutes. Positive and negative controls were tested in replicates and triplicates subsequently. Following incubation, the wells’ contents were discarded and the plates were washed for five times as mentioned above. Diluted in a dilution buffer, anti-human IgG horseradish peroxidase (HRP) conjugate was applied to plates (100 μl / well) that were incubated at 37 °C for 10 minutes. Then the plates were washed for five times as before and TMB substrate solution was added (100 μl / well), followed by incubation for 5 minutes at room temperature at dark. By adding ELISA stop solution (100 μl / well), the colour development was disrupted. The optical density (OD) had been measured at 450nm.

### Determination of the cut-off value

The cut off value was determined with the negative samples. The mean OD of the negative controls has been determined and a sample is considered positive when the sample OD value at 450 nm exceeds the mean OD value of negative controls plus three times the standard deviation (SD), denoting as cut-off value. For a sample to be negative the value should be equal or less than the cut-off OD.

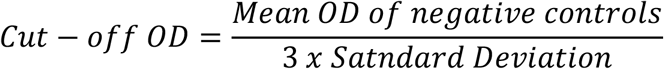

### Internal validation of assay

The validation of the assay was evaluated on the basis of four critical aspects such as the form and the number of specimens, identification and visualization of reagents, and the overall time of the assay.

### Reproducibility

Reproducibility of developed assay was determined by intra and inter assay. Five replicates of positive and negative sera on the same plate within a day was analysed to determine intra-assay variance. And the inter-assay variation was observed on 05 separate working days using positive and negative controls and the coefficient of variation was calculated using the following formula,

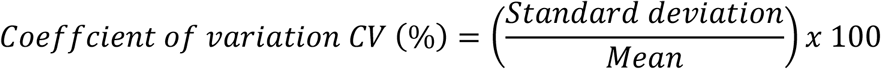

### Clinical Validation

In order to define clinical sensitivity and specificity, both positive and negative panels were assayed with the developed ELISA systems.

### Ethical Statement

This study was approved by National Research Ethics Committee (NREC) of Bangladesh. Well informed consents were obtained from all the participants. During the collection of samples, the objectives of the study and information on use of data were provided to potential participants. Participants were allowed to ask questions and were to seek clarification about the study.

### Statistical Analysis

Statistical analysis was performed using STATA 13 (StataCorp, LP, College Station, Texas, USA) and the graphical presentation was made by using GraphPad Prism (7.05). A p-value of <0.05 was considered as significant. Sensitivity, specificity, positive predictive value and negative predictive value and area under curve (AOC) with 95% confidence interval characterized the feasibility of these ELISA assays was performed between the true positive and in-house ELISA methods. Significant association between the in-house developed IgG ELISA assay for NCP, S1 and RBD antigens was analysed using the same sample panels. Cohen’s Kappa test was used to estimate the test agreement.

## Result

### Optimization and standardization of ELISA Assay

Using checkerboard method, the assay condition that exhibited higher signal to noise ratio with acceptable background have been selected separately for either S1 or RBD (Table 1 and 2). S1 protein of 0.40 µg/well, with 1:4000 dilution of HRP-conjugated secondary antibody and 1:100 diluted sera were the optimal variables selected for system 1. For system 2, the assay was optimized with coating concentration of 0.20 µg/well for sample diluted at 1:100 and 1:4000 diluted secondary antibody. The incubation time for both of the assays was set as 30 minutes.

**Table 1:**
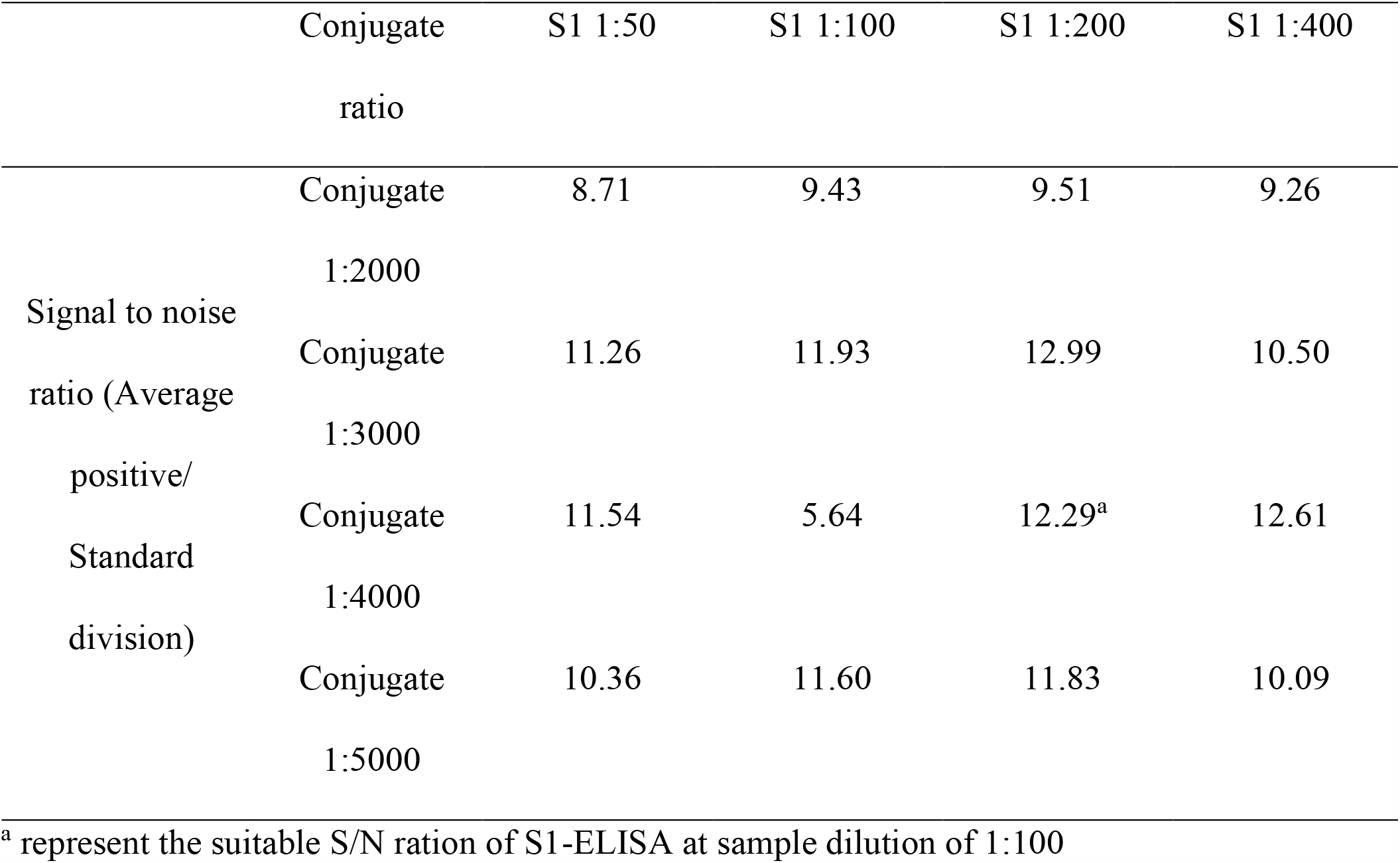
Checkerboard assay for S1-ELISA using positive samples

**Table 2:** Checkerboard assay for RBD-ELISA using positive samples

|  | Conjugate | RBD 1:50 | RBD 1:100 | RBD 1:200 | RBD 1:400 |
| --- | --- | --- | --- | --- | --- |
|  | ratio |  |  |  |  |
|  | Conjugate | 7.31 | 7.13 | 9.46 | 10.36 |
| Signal to | 1:2000 |  |  |  |  |
| noise ratio | Conjugate | 9.57 | 10.01 | 11.30 | 2.24 |
| (Average | 1:3000 |  |  |  |  |
| positive / | Conjugate | 10.73 | 11.14 | 11.57<sup>a</sup> | 11.75 |
| Standard | 1:4000 |  |  |  |  |
| division) | Conjugate | 10.24 | 10.57 | 10.78 | 10.38 |
|  | 1:5000 |  |  |  |  |

<sup>a</sup> represent the suitable S/N ration of RBD-ELISA at sample dilution of 1:100

### Performance evaluation

#### Reproducibility and precision

The intra-assay and inta-assay variation were determined and analysis showed that for intra-assay coefficient of variance is <10% for system 1 and <15% for system 2, whereas for inter-assay the variance is <25% for both the assays (Table 3 and 4).

**Table 3:** Reproducibility and Precision of the in-house S1 specific ELISA (System 1)

| Controls used | CV (inter assay) | CV (intra assay) |
| --- | --- | --- |
| Positive control P1 | 18.03 | 6.64 |
| Positive control P2 | 22.61 | 5.29 |
| Negative control N1 | 21.09 | 3.25 |
| Negative control N2 | 19.76 | 7.31 |

**Table 4:** Reproducibility and Precision of the in-house RBD specific ELISA (System 2)

| Controls used | CV (inter assay) | CV (intra assay) |
| --- | --- | --- |
| Positive control P1 | 22.98 | 6.09 |
| Positive control P2 | 21.66 | 3.81 |
| Negative control N1 | 24.20 | 2.68 |
| Negative control N2 | 21.39 | 15.00 |

#### Clinical validation

For performance validation, the sensitivity of the systems has been determined with panel 1 and panel 2 comprised of nineteen and sixty samples respectively that were previously studied [21].

The mean OD/cut-off ration in system 1, for panel 1, panel 2, healthy negative control and pre-pandemic dengue sample were (2.53±2.01), (7.55±4.04), (0.58±0.11), and (0.65±0.14) respectively (Figure 1B). Analysis showed that system 1 with S1 as immobilizing agent, can successfully detect 73.68% (95% CI: 48.8%, 90.9%) of true cases with onset of symptoms <14 days, with test agreement 83.0% (Kappa=0.83, p<0.001) (Table-5). The positive predictive value (PPV) and negative value (NPV) were 93.1% and 95.5%, respectively (Table 6).

**Fig 1.**
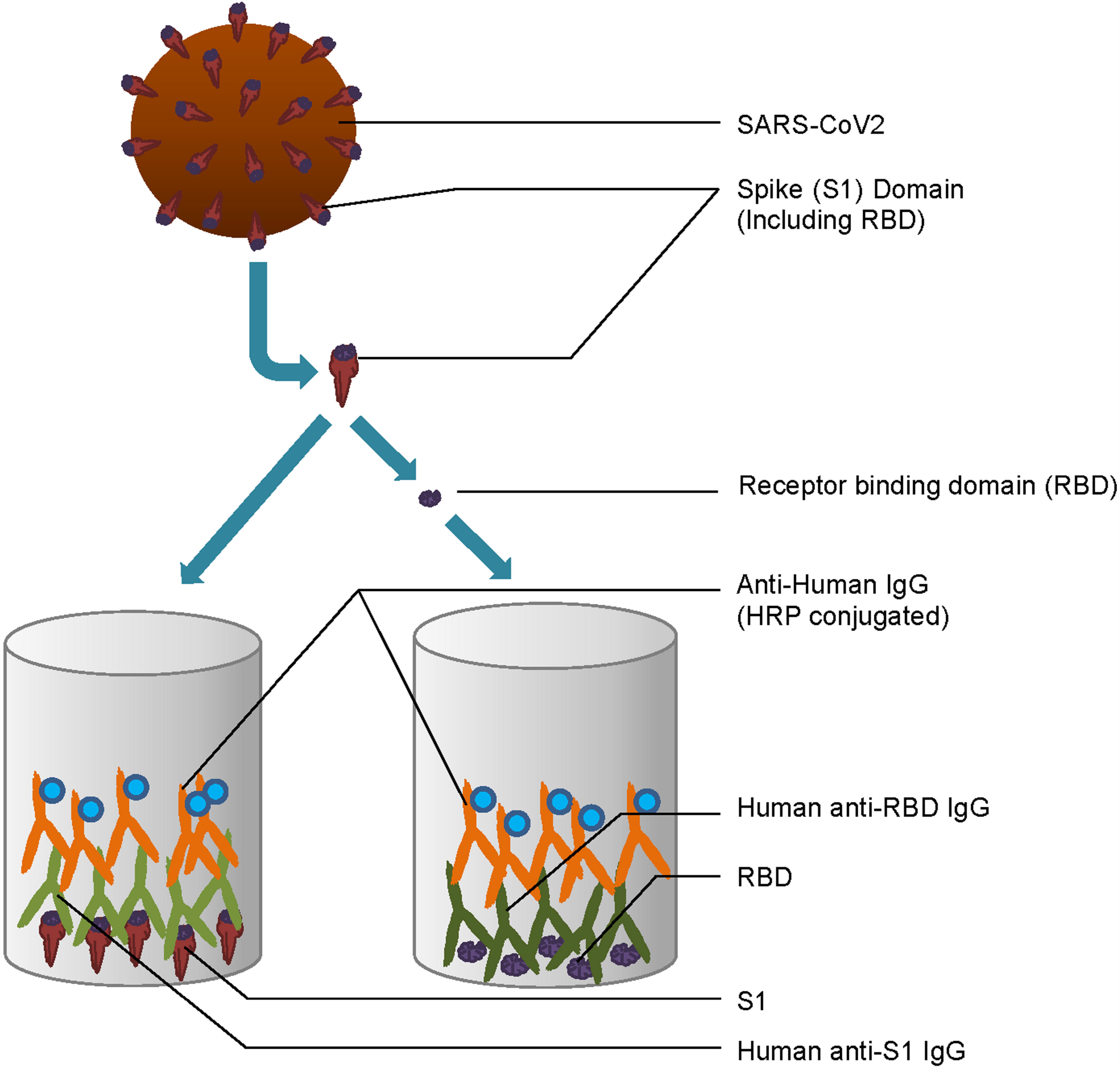
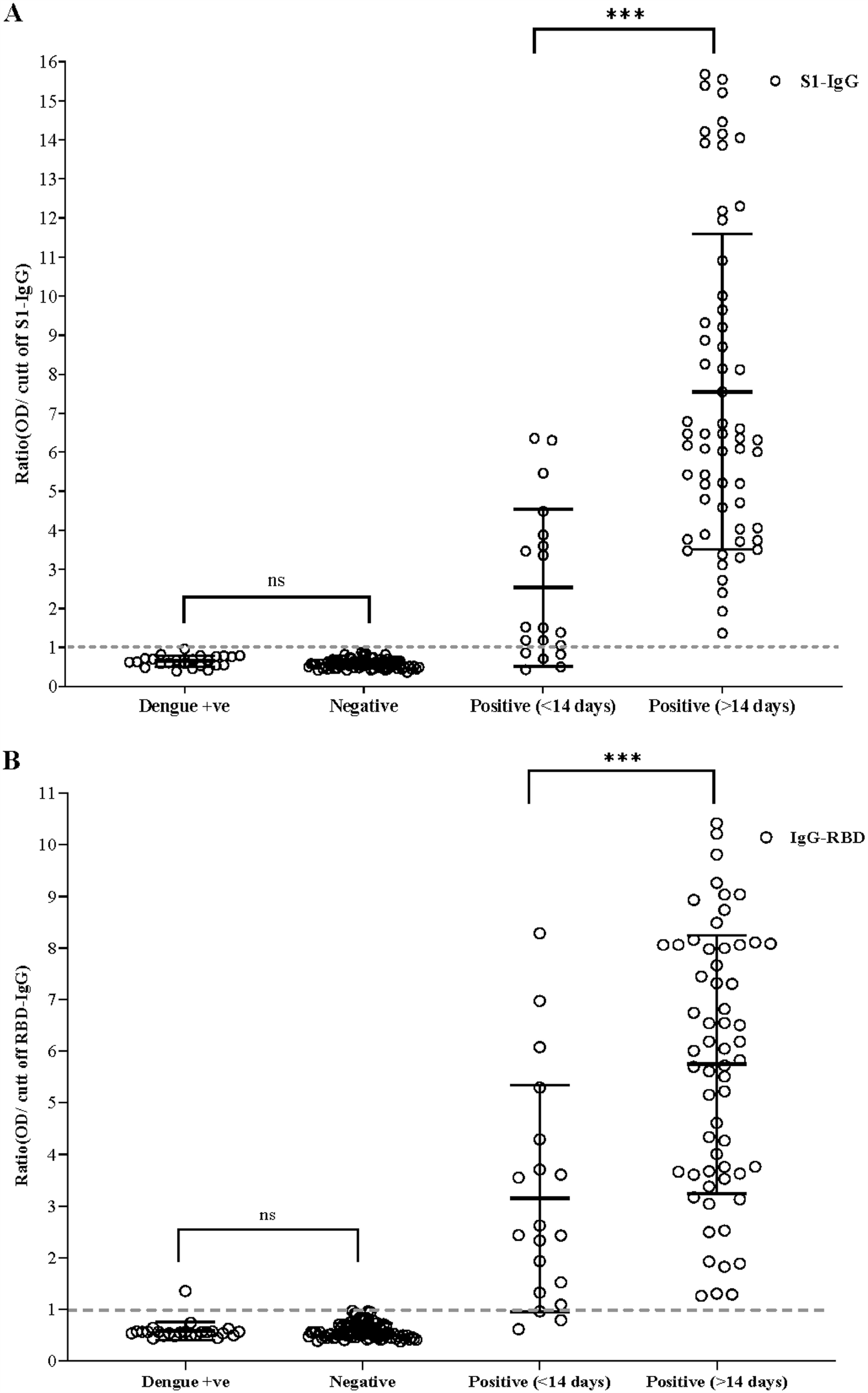
Detection of SARS-CoV2 RBD-IgG (A) and S1-IgG among healthy donors, dengue positive samples, and SARS-CoV2 confirmed patients. Ratio of OD/Cut off RBD-IgG and S1-IgG value of negative, dengue positive and positive with SARS-CoV-2 within 14 days and more than 14 days were shown. Data are presented as mean with ± Standard deviation. The p-value was calculated by independent sample t-test. The reference line indicating the cut-off of the in-house ELISA methods.

Sensitivity reaches its peak for samples collected after >14 days of disease onset detecting all of the cases, i.e. 100% (95% CI: 94.0%, 100%), with test agreement 100.0% (Kappa=1.00, p<0.001). Interestingly, both the PPV and NPV was 100%. The overall, sensitivity and specificity of the assay were 94.9 (95% CI: 87.4%, 98.6%) and 100 (95% CI: 96.5%, 100%), respectively with strong Kappa agreement of 94.0% (p<0.001) (Table 5 and 6). Overall, PPV and NPV for S1-ELISA were 100% and 96.3%, respectively.

**Table 5:** Sensitivity, specificity and area under curve (AUC) analysis for RBD and S1 antigen against IgG in RT-PCR positive against SARS-CoV-2 and pre-pandemic and healthy controls.

| Protein | Days | Sensitivity, %<br>(95% CI) | Specificity, %<br>(95% CI) | AUC<br>(95% CI) | Test<br>agreement | p-value |
| --- | --- | --- | --- | --- | --- | --- |
| S1 | ≤ 14 | 73.7<br>(48.8, 90.9) | 100<br>(96.5, 100) | 0.87<br>(0.77, 0.97) | 83.0% | <0.001 |
|  | > 14 | 100<br>(94.0, 100) | 100<br>(96.5, 100) | 1.0<br>(1.0, 1.0) | 100% | <0.001 |
|  | Overall | 94.9<br>(87.4, 98.6) | 100<br>(96.5, 100) | 0.97<br>(0.95, 1.00) | 94.0% | <0.001 |
| RBD | ≤ 14 | 84.2<br>(60.4, 96.6) | 99.0<br>(94.8, 100) | 0.92<br>(0.83, 1.0) | 87.0% | <0.001 |
|  | > 14 | 100<br>(94.0, 100) | 99.0<br>(94.8, 100) | 1.0<br>(0.99, 1.0) | 99.0% | <0.001 |
|  | Overall | 96.2<br>(89.3,99.2) | 99.0<br>(94.8,100) | 0.98<br>(0.95,100) | 96.0% | <0.001 |

Note; RBD, Receptor Binding Domain; S1, Spike-1; CI, Confidence Interval; AUC, Area Under curve

**Table 6:** Positive and Negative predicted value of RBD and S1 antigen against IgG in RT-PCR positive against SARS-CoV-2 and pre-pandemic and healthy controls.

| Protein | Days | PPV<br>(95% CI) | NPV<br>(95% CI) |
| --- | --- | --- | --- |
| S1 | $\leq 14$ | 93.1 | 95.5 |
|  |  | (76.8, 100) | (89.7, 98.5) |
| | $> 14$ | 100 | 100 |
|  |  | (94.0, 100) | (96.5, 100) |
|  | Overall | 100 | 96.3 |
|  |  | (95.1, 100) | (90.9, 99.0) |
| RBD | $\leq 14$ | 94.1 | 92.2 |
|  |  | (71.3, 99.9) | (92.0, 99.4) |
| | $> 14$ | 98.4 | 100 |
|  |  | (91.2, 100) | (96.5, 100) |
|  | Overall | 98.7 | 97.2 |
|  |  | (93.0, 100) | (92.0, 99.4) |

Note; RBD, Receptor Binding Domain; S1, Spike-1; CI, Confidence Interval; PPV, Positive predicted value; NPV, Negative predicted value

The mean OD/cut-off ratio in system 2, for panel 1, panel 2, healthy negative control and pre-pandemic dengue sample were (3.15±2.19), (5.74±2.50), (0.58±0.15), and (0.58±0.18), respectively (Figure 1B). For RBD-specific IgG ELISA, the sensitivity was higher than that of S1 during the early phase of infection i.e. for panel 1 samples; the sensitivity value was 84.21% (95% CI: 60.4%, 96.6%) with strong test agreement 87.0% (Kappa=0.87, p<0.001), which increased to 100% (95% CI: 94%, 100%) for panel 2 containing samples obtained after 14 days of onset of symptoms, with test agreement 99.0% (Kappa=0.99, p<0.001). Overall sensitivity was 96.2 (95% CI: 89.3%,99.2%), relatively better one than S1-ELISA (Table-5), though the specificity slightly reduced to 99.0% (95% CI: 94.8%,100%) with kappa test agreement of 96% (p<0.001). PPV for pane1, panel 2 and overall assay were 94.1%, 98.4% and 98.7%, respectively, whereas, NPV for those panels were 92.2%, 100% and 96.3%, respectively.

### Comparative analysis of RBD and S1 ELISA with NCP ELISA

When comparative analysis were performed with the three in-house ELISA systems using the same panels, the mean OD/cut-off ratio for NCP-, S1- and RBD-ELISA for samples collected within 14 days of onset of symptoms were (mean ± std) (3.22±2.55), (3.15±2.19), (2.53 ±2.01), respectively, which were (4.24±2.55), (5.12 ±2.66) and (6.34±4.24) for sample collected after 14 days of onset of symptoms. However, the overall mean OD/cut-off ratio for these three assays were (4.56±2.48), (5.74±2.50), and (7.55±4.04), respectively (Figure 1A, 1B). The overall mean of OD/cut-off ratio for these assays were (Figure 1C). Moreover, NCP-IgG ELISA strongly correlated with RBD-IgG (r_s_=0.80; p<0.001) and S1-IgG (r_s_=0.74; p<0.001) ELISA systems. Similar association was noted between the RBD-IgG and S1-IgG (r_s_=0.804; p<0.001) ELISA systems (Figure 3A, 3B, 3C).

**Fig 2.**
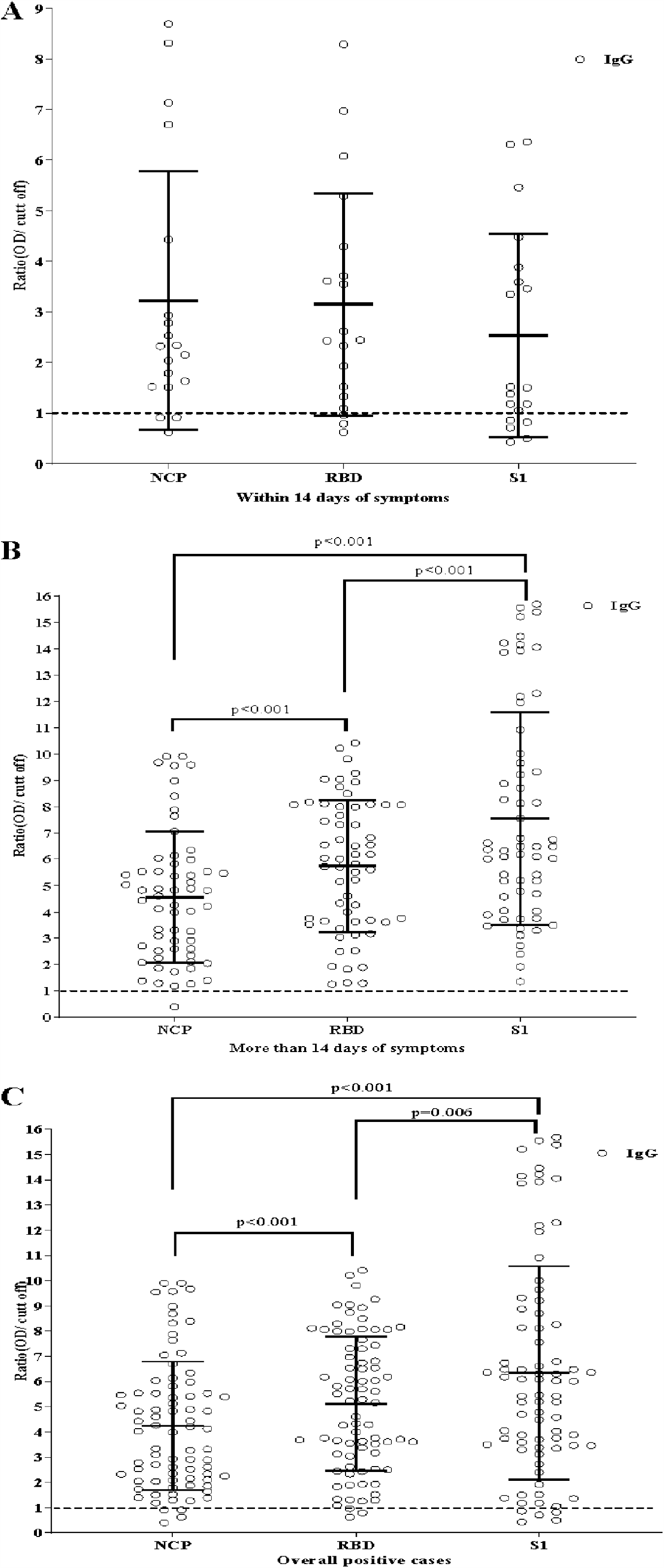
Detection of SARS-CoV2 NCP-IgG, RBD-IgG and S1-IgG among the SARS-CoV2 confirmed patients with overall period (A) within 14 days (≤ 14) (B) and more than 14 days (> 14) (C). Ratio of OD/Cut off of NCP-IgG, RBD-IgG and S1-IgG of the confirmed positive with SARS-CoV-2 were shown. Data are presented as mean with ± Standard deviation. The p-value was calculated by independent sample t-test. The reference line indicating the cut-off of the in-house ELISA methods.

**Fig 3.**
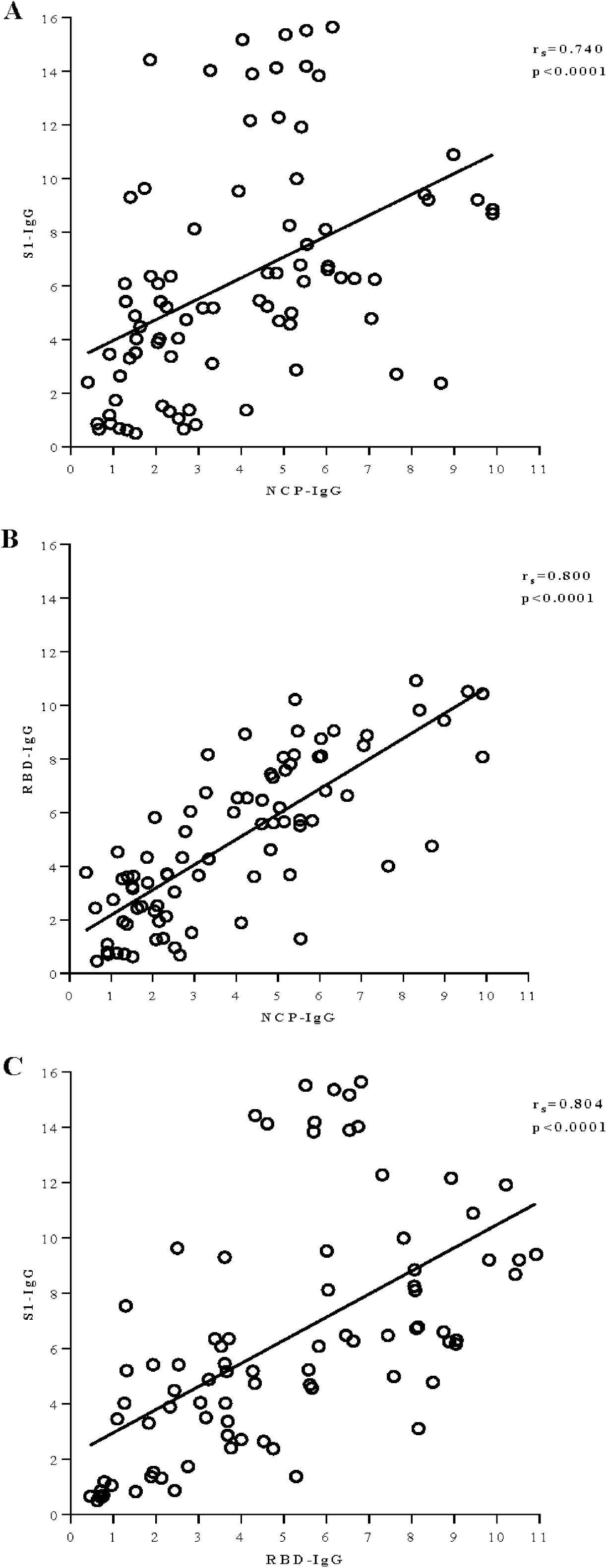
Correlation between NCP-IgG and RBD-IgG (A), NCP-IgG and S1-IgG (B) and RBD-IgG and S1-IgG were shown. Spearman rank correlation was used to estimate the p-value. A p-value of <0.05 was considered as significant.

## Discussion

The disadvantage of molecular diagnostics i.e. viral RNA detection, in later phase diagnosis of COVID-19 infection, necessitates the implementation serological tests to better profile the disease outcome [22]. Scientists across the world are now contemplating on the development of several immunoassays resulting in the currently available test procedures based on either Enzyme Linked immunosorbent Assay (ELISA), Chemiluminescent assay (CLIA), Lateral Flow Immunoassay (LFIA), etc. [20]. However, a meta-analysis and system review of currently available serological assays, reported lower sensitivities specially for LFIA and resulted in 18% of COVID-19 patients being misdiagnosed as negative by IgG ELISA when tested after 21 days of onset of symptoms [23]. The surging demand of the affected countries of serological tests has outpaced the development and production of efficient and accurate immunoassays, irrespective of the relaxation in the tedious and rigorous evaluation.

Countries’ decision-making such as providing “immunity-passport” as well as resuming normal activities require tracking of individuals with neutralizing antibodies which may provide immunity against reinfection [24, 25]. S1 subunit of spike protein and S-RBD play a vital role in binding with human Angiotensin Binding Receptor-2 (hACE-2) initiating a cascade of events leading to viral entry. Residues 445-456, 473-477, and 484-505 of RBD are found to interact with hACE-2 [26, 27]. Therefore antibodies to either of the three epitopes of RBD or epitopes of S1 can interfere with the binding step contributing to viral neutralization, thereby rendering anti-RBD IgG titre as a prediction for protection and survivability [28-30]. In an investigation anti-RBD IgG was found to exist for up to 75 days from presentation of COVID-19 disease with no cross reaction with other known circulating human coronaviruses [31, 32]. Hence, ELISA assay with immobilized RBD or S1 protein can be a plausible tool for implementing detection of seroconversion with protective immunity and evaluating vaccine efficacy.

This study demonstrates the development of indigenous ELISA assays targeting antibodies against spike S1 protein and receptor binding domain (RBD). To our knowledge this report is the first to address ELISA assays with 30 minutes of assay time, specific for S1-IgG and RBD IgG. Our study corresponds the overall sensitivity of in-house RBD-ELISA and S1-ELISA to be 96% and 95% respectively with slightly higher positive predictive value for S1 (Table 5 and 6). However, about 84% of the RT-PCR positive samples collected within two weeks of symptoms appearance, showed positive reaction in RBD-ELISA reaching a peak of 100% in RT-PCR positive samples with >14 days of COVID-19 symptom onset (Figure 1A, 1B; Table 5). This relation between antibody appearance and duration of onset of disease was also established by other studies [30, 33]. However, for S1 as coated antigen, about 74% of the former group exhibited positive serology though for the latter, the sensitivity is indifferent (Table 5). Therefore, ELISA system targeting anti-RBD IgG is more sensitive than anti-S1 ELISA during the early phase of infection (Figure 1A, 1B; Table 5 and 6).

Antibodies against RBD were detected in 1% of the healthy donor and pre-pandemic negative sera, whereas the positivity is nullified for these negative sera when tested with S1-ELISA system (Table 5). Our findings were similar to a study where 1.9% sera of pre-pandemic and healthy donors were found positive for SARS-CoV-2 cross-reacting antibodies [33].

Antibodies against N protein detected by inhouse-IgG ELISA can be compared with those with inhouse S1 and RBD ELISA [21]. Comparing to the previous study, it is observed that the sensitivity of nucleocapsid specific IgG ELISA and RBD-ELISA are equal for early phase samples and higher than S1-ELISA (Figure 2A). However, for samples collected after two weeks from symptom onset, NCP specific IgG ELISA misclassifies 2% of the positive samples as negative (98% sensitivity) (Figure 2B), which was also observed by others [34]. Our finding is in contrast with studies that report higher prevalence of N targeted IgG in comparison with S1-RBD [16], but coincide with [19], where overall antibody titre against S1 was found higher than NCP (Figure 2C). The possible explanation behind higher level and primordial inception of IgG against NCP may be 90% amino acid homology and previous exposure to SARS or other human coronaviruses leading to plausible cross-reaction and decline in specificity [35, 36]. For RBD, the amino acid homology between SARS-COV and SARS-COV-2 has been reported to be 72% and cross-reactivity to a lesser extent among antibodies [37]. Also monoclonal antibody specific for SARS-CoV RBD has been isolated have binding capacity with an epitope of SARS-CoV-2 RBD which is free from binding with hACE-2 [27]. Heterogeneity in dynamics and kinetics of antibodies specific to different antigens have also been reported [33]. Our study reports that overall correlation among the three in-house ELISA systems is significant (Figure 3A, 3B, 3C).

There are certain limitations of our study that are to be addressed. Cross reactivity against SARS-CoV and other coronaviruses could not be assessed, and the size of sample collected within 14 days of symptom onset is relatively small. Nonetheless, the developed ELISA assays can be implemented in dengue-endemic countries due to their least cross reactivity and the assays are free from spectrum biasness.

From our findings, we recommend the use of NCP and RBD Ig-ELISA kit for early detection studies of SARS-CoV-2 antibodies, with former being the better choice because of higher specificity. Whereas for retrospective sousveillance both S1- and RBD-ELISA can be implemented, with former being choice of assay for low prevalent areas. However, future endeavours should include longitudinal study with large cohort of population to access the antibody dynamics and persistence for the effective implementation of vaccine and eradication of COVID-19 from the population.

## Data Availability

All data underlying the findings in our study are freely available in the manuscript. For additional information please refer to: http://www.grblbd.com

## Author Contribution

Nihad Adnan, Bijon Kumar S. and Mohd. Raeed J. designed and supervised this research. Mumtarin Jannat O., Nowshin Jahan., Shahad Sharif K., Tammana Ali. performed the experiment. Md. Ahsanul H. performed the data curation and statistical analysis. Mohib Ullah K., Eiry Kobatake, and Masayasu Mie proof read it. All authors contributed to the construction and writing this article.

## Acknowledgement

The authors express their thanks all the participants who were willing participate in the study. They also thanks their team members for their support work during the current pandemic situation.

## Ethical Conduct of Research

Ethical approval for this study was issued by the National Research Ethics Committee (NREC) of Bangladesh. Approval number BMRC/NREC/2019-2022/697.

